# The hidden productivity toll of perimenopause: symptom-driven work impairment during women’s peak earning years

**DOI:** 10.64898/2026.07.05.26357306

**Authors:** Yihan Xu, Carley Prentice, Yella Hewings-Martin, Adam Cunningham, Liudmila Zhaunova, Jaume Puig-Junoy

**Author notes:** Correspondence should be addressed to Yihan Xu, 27 Old Gloucester Street, London, WC1N 3AX, United Kingdom.

## Abstract

Perimenopausal women, often in the peak-earning years of their careers, make up a significant proportion of the workforce, with women aged 40-49 accounting for approximately 21% of the U.S. female workforce. Previous studies have revealed perimenopause to be associated with psychological, vasomotor, somatic, and cognitive symptoms that affect women’s quality of life, yet its workplace and economic consequences remain under-explored. We examined work impairment by symptom severity and across reproductive stages in a cross-sectional survey of U.S. women aged 35-59 (n=945), using the Work Productivity and Activity Impairment questionnaire and Menopause Rating Scale. We then estimated associated productivity losses using a human capital approach.

Perimenopausal women were equally likely to remain in the labour force as premenopausal women (76.6% vs. 78.0%) but reported substantially higher work impairment (22.5% vs. 12.7%). Work impairment rose from 3.4% among women with minimal symptoms to 33.4% among those with severe symptoms and was driven predominantly by presenteeism rather than absenteeism. Somatic and psychological symptoms showed the strongest associations with work impairment, whereas urogenital symptoms were not significantly associated. The observed work impairment translated into an estimated annual productivity loss of approximately $6,061 per woman in perimenopause and a societal burden of $56.7 billion in the United States. These findings suggest that perimenopause is a substantial but under-recognised workplace health challenge that requires tailored, symptom-matched, workforce support.

## Introduction

Perimenopause, the transition phase preceding confirmed menopause, is a common yet under-explored stage of women’s lives ^1^. It lasts around 4-8 years with a typical onset at 45 years old ^2,3^. The transition is characterised by fluctuating vasomotor, psychological, and somatic symptoms that can significantly affect women’s quality of life, daily functioning, and productivity ^4,5^. Despite the universality of this life stage and the prevalence of the symptoms ^6^, perimenopause remains under-recognised by women and clinicians ^7,8^. About one in three women above 35 years in the United States report uncertainty about perimenopause ^9,10^ and frequently mis-attribute symptoms arising from hormonal changes (e.g. exhaustion, sleep difficulties, joint and muscle pains) to stress, ageing, or competing life demands ^8,11^. While the effects of other major reproductive-health related experiences (menstruation, motherhood, and menopause) on women’s working lives have received growing attention ^12–15^, perimenopause remains an under-recognised and under-researched workplace challenge.

This lack of recognition may carry important economic consequences as perimenopausal women make up a significant portion of the workforce, with women aged 40-49 accounting for approximately 21% of the U.S. female workforce ^16^, and are often in the peak-earning years of their careers ^13,17^. Although emerging studies have linked symptoms during menopausal transition to workplace productivity, much of the evidence has focused on menopause or vasomotor symptoms ^12,18,19^. Perimenopause, however, may pose unique challenges, for it is the earlier, more uncertain phase of menopausal transition, when symptoms may first appear, fluctuate, intensify, and become disruptive, often following highly individual trajectories ^10,20,21^. At this stage, women often do not yet recognise them as hormonally related or seek support, because the initial symptoms are more likely to arise in the mental health, sleep, and cognitive domains rather than the more classical vasomotor symptoms that typically appear in later transition ^22,23^. Delays in symptom recognition and support may result in long-term, unaddressed work impairment, creating opportunity costs through reduced productivity, slower career progression, and potentially spillover effects extending into menopause ^12^. As a result, the perimenopause-related workplace productivity loss may be substantial yet systematically overlooked.

Another challenge in measuring the workplace impact of perimenopause is unpacking how different symptom domains impair functioning, as they may do so via distinct mechanisms. For example, sleep disruption and musculoskeletal pain may reduce physical endurance, alertness, and attention ^24,25^, anxiety and mood disturbance may impair emotional regulation and interpersonal functioning ^26^, whereas cognitive symptoms may affect concentration, memory, and decision-making ^27^. Yet most existing studies have treated menopausal symptoms as a uniform construct or focused narrowly on vasomotor symptoms ^28^. This approach may mask important differences in how symptoms affect work impairment during perimenopause, limiting our ability to identify the support individual employees need and to design workplace policies responsive to different forms of symptom-related impairment. Overall, there is scant evidence on the impact of perimenopause on women in the workplace and its broader economic consequences, and the limited understanding of the mechanisms of how symptoms drive work impairment. Therefore, this study aims to examine work impairment across reproductive stages and symptom severity, identify the symptom domains most strongly associated with work impairment, and estimate the productivity loss associated with perimenopause in the United States.

## Methods

### Study design and ethics

We conducted a cross-sectional study among U.S. women aged 35 to 59 years old. We collected data between February 27 and March 16, 2025, using SurveyMonkey, a digital survey platform. The Western Copernicus Group Institutional Review Board approved this study (IRB No. 20250554).

### Participants and recruitment

Participants were recruited via Prolific, an online research participant recruitment platform, using pre-defined eligibility criteria (see Figure 1 for participant flow diagram). Potential participants first completed a screener on Prolific, which was restricted to individuals residing in the United States, aged 35-59 years, of female sex, and fluent in English. We also set age quotas to approximate the distribution of women likely to be in perimenopause based on prevalence estimates from a prior study ^22^. Participants provided e-consent to take part in the study and were excluded if they had conditions or circumstances likely to confound the assessment or experience of perimenopausal symptoms or their impact on work productivity, including current pregnancy, recent birth, recent pregnancy loss, or a history of hysterectomy. Participants who did not provide reproductive stage information were also excluded from the final study sample. Participants were remunerated at $12–15 USD per hour. Survey responses were collected anonymously.

**Figure 1.**
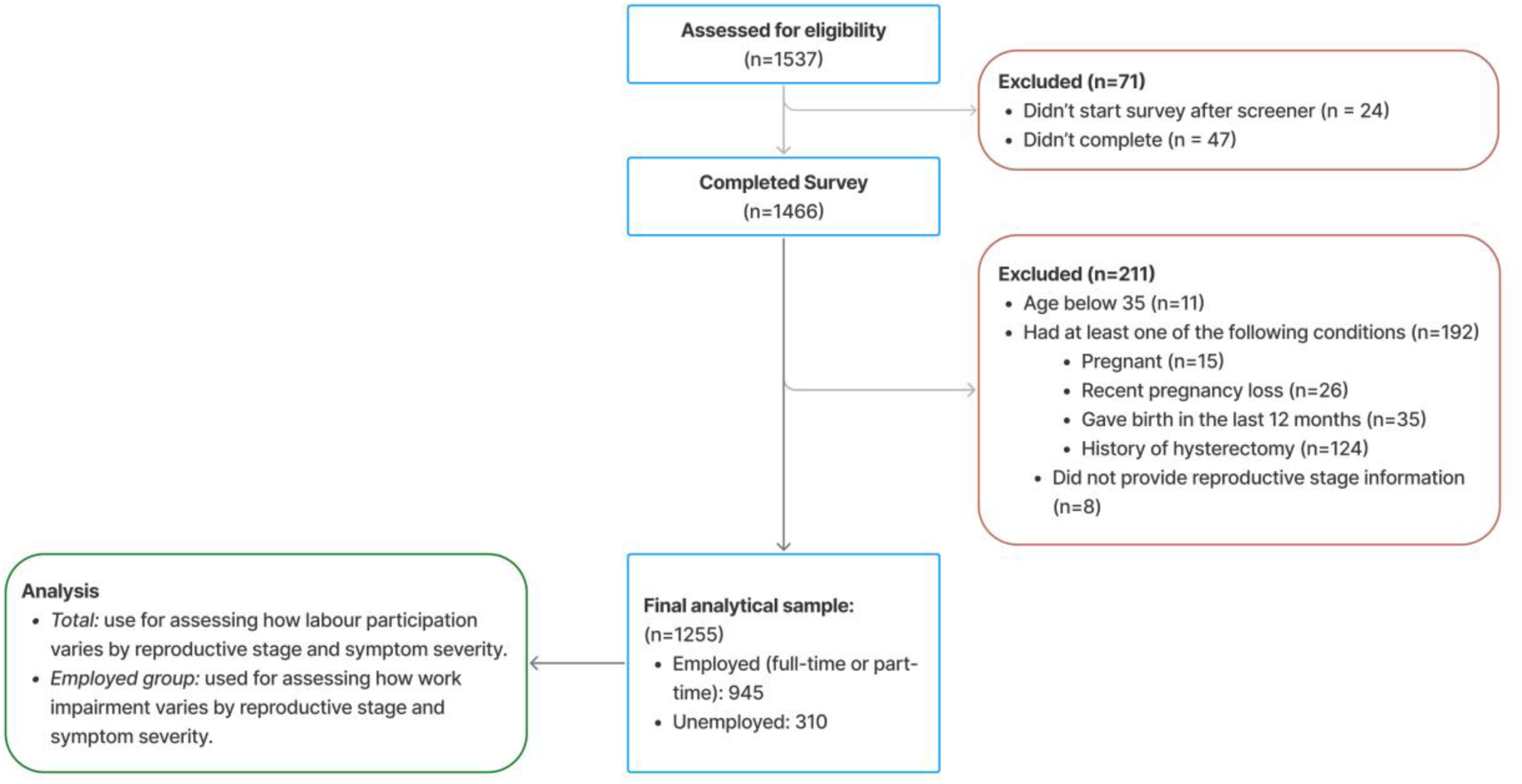
Participant flow.

### Measures

#### Work impairment

Work impairment was measured using the Work Productivity and Activity Impairment questionnaire (WPAI), one of the most commonly used instruments in health economics evaluations ^29,30^. Before the relevant WPAI items, participants in paid employment were explicitly asked to “consider **only** how much perimenopause symptoms affected” their productivity or regular daily activities to minimize confounding effects from symptoms from other health conditions or life circumstances. Work impairment comprises absenteeism (percentage of work time missed due to symptoms) and presenteeism (reduced productivity while working) during the preceding seven days; it ranges from 0% to 100%, with a higher percentage indicating greater impairment. While the WPAI also assesses impairment in non-work activities, this study used the overall work impairment as the primary productivity outcome, which formed the basis for estimating productivity loss in the subsequent economic impact evaluation section.

#### Health-related quality of life

To capture broader health-related outcomes beyond work impairment, we chose EuroQol five-dimensional (EQ-5D-5L) as a secondary outcome. Participants completed the EQ-5D-5L questionnaire, and their health-related quality of life utility scores were derived using the U.S. value sets. The theoretical values range from −0.573 to 1, where 1 represents full health, 0 represents dead, and negative values indicate health states valued as worse than dead ^31,32^.

#### Symptom severity

Symptom severity was measured by the Menopause Rating Scale (MRS), a widely used validated scale for women navigating the menopausal transition ^33^. The MRS is an 11-item questionnaire that measures severity of symptoms, which fall into three distinctive domains: somatic, psychological, and urogenital. The somatic domain comprises hot flashes/sweating, heart discomfort, sleep problems, and joint/muscular discomfort; the psychological domain comprises depressive mood, irritability, anxiety, and physical/mental exhaustion; and the urogenital domain comprises sexual problems, bladder problems, and vaginal dryness. Each item is scored on a 5-point Likert scale (0 = none, 4 = very severe), with higher scores indicating greater symptom burden. Both domain scores and total scores were calculated by summing up individual scores of relevant items. In this study, symptom burden was analysed both categorically based on defined thresholds (minimal: 0-4, mild: 5-8, moderate: 9-16, and severe: ≥17) and continuously when exploring how domain scores were associated with work impairment. In addition, the symptom severity analyses included participants across reproductive stages (premenopausal, perimenopausal, postmenopausal, and those who were unsure of their stage).

#### Reproductive stage

Participants self-identified their reproductive stage using a single-item question adapted from prior research ^10^. After reviewing descriptions and a visual representation of different reproductive stages (Supplementary Material A), participants were asked to select the stage that best described them: premenopausal, perimenopausal, postmenopausal, or unsure.

#### Demographics

Demographics variables, including education, ethnicity, height, and weight were self-reported. Participants reported their highest level of education completed using predefined response options, which were subsequently grouped as less than high school, high school graduate, some college or associate degree, bachelor’s degree, and graduate or professional degree. Ethnicity was assessed using a multi-select question, with responses classified for analysis as White, Asian, Black, Hispanic, Mixed, or Other. Participants reported their current height in either feet and inches or centimeters and their current weight in either pounds or kilograms. Body Mass Index (BMI) was subsequently derived from height and weight and classified as underweight (<18.5 kg/m^2^), normal weight (18.5-24.9 kg/m^2^), overweight (25.0-29.9 kg/m^2^), or obesity (≥ 30 kg/m^2^).

### Statistical analysis

Descriptive statistics were used to summarise participant characteristics and distributions of outcome measures. Differences in work impairment across reproductive stage and symptom severity groups were assessed using Kruskal-Wallis tests due to the non-normal distribution of WPAI outcomes.

Multivariable linear regression models were used to examine how work impairment and EQ-5D utility differ by reproductive stage and symptom burden. Reproductive stage and symptom severity were examined in separate multivariable models. Models assessing reproductive stage were adjusted for age group, BMI, education, and ethnicity. Models assessing symptom severity were adjusted for the same covariates. Reproductive stage and symptom severity were not mutually adjusted because symptom severity may lie on the pathway between reproductive stage and work impairment or health-related quality of life; mutual adjustment would therefore estimate a conditional rather than an overall association. Regression results are reported as β coefficients with 95% confidence intervals (CIs).

All analyses were conducted in R (version 4.4.1), with statistical significance level at p = 0.05.

### Economic impact evaluation

Productivity loss, the economic outcome, was estimated by converting work impairment into monetary losses using the human capital approach, which values healthy time using market wage rates, such that work impairment is translated into monetary losses based on typical earnings ^34,35^.

At the individual level, productivity loss was estimated by converting differences in work impairment into lost earnings. Conceptually, productivity loss can be expressed as:

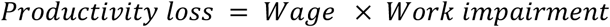

Where work impairment represents the proportion of working time lost due to either absenteeism or presenteeism, as measured by the WPAI instrument.

The wage estimates were sourced from the median weekly earnings for U.S. women according to the U.S. Bureau of Labor Statistics ^16^. For individual-level estimates, the median weekly wage corresponding to women aged 45-54 was applied and annualised assuming 52 working weeks per year, whereas age-group specific wages were applied for population-level estimates.

Perimenopause-associated work impairment was calculated as the difference in mean work impairment (from 0 to 100%) between the premenopausal and perimenopausal participants. Similarly, work impairment associated with symptom burden (as measured by the MRS) was calculated as the difference in mean work impairment between participants with minimal symptom level and those with mild, moderate, and severe symptoms. Therefore, the productivity-loss estimates should be interpreted as incremental losses relative to an empirical reference rather than relative to an assumption of 100% productivity.

Observed (descriptive) differences are reported in main results to describe the raw magnitude of work impairment across reproductive stages and symptom severity, whereas regression-adjusted differences are provided in the Supplementary Materials (see Supplementary Table 2a, 2b and 3) to support interpretation of the association between perimenopause and work impairment.

At the population level, total work impairment associated with perimenopause was estimated by applying individual-level perimenopause-related work impairment to the U.S. female population aged 35–59, accounting for age-specific population size, labour force participation rate, and the estimated prevalence of perimenopause within each age group. Conceptually, it can be expressed in the formula below:

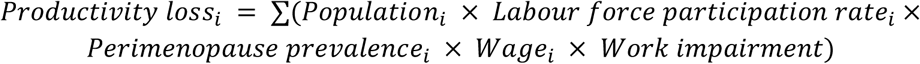

Where *i* indexes age groups; Populationᵢ denotes the number of women in each age group; Labour force participationᵢ represents the proportion of women in the workforce; Perimenopause prevalenceᵢ is the estimated proportion of women in perimenopause; and Wageᵢ corresponds to age-specific median weekly earnings. Work impairment reflects the proportion of productive time lost, as defined above.

Age-group specific female population, weekly median earnings, and labour force participation rates were obtained from publicly available U.S. statistics ^16,36,37^. Age-group specific prevalence of perimenopause was modelled based on published epidemiological data on the age distribution of menopausal transition ^2,38^. The onset of perimenopause was assumed to follow a right-skewed distribution centered around a mean age of 45 years with a standard deviation of 4.5 years, reflecting the heterogeneity of timing on the onset. As the range of perimenopause was commonly reported to be 4 to 8 years, a typical duration of 6 years was assumed to approximate the proportion of women in perimenopause within each age group.

All monetary estimates are presented in 2025 U.S. dollars.

## Results

### Participant characteristics

A total of 1,255 women aged 35 to 59 were included in the study (mean age 45.5 years, SD 5.4), of whom 50.5% self-identified as perimenopausal. Of these, 945 participants were in paid employment (full-time or part-time) and constituted the analytic sample for analyses of work productivity. Figure 1 presents the eligibility criteria and participant flow.

Table 1 presents the demographic characteristics for the employed participants by self-reported reproductive stage. This sample was predominantly White (72.5%), with similar education levels and BMI. More than half of participants self-reported as in perimenopause (51.3%), and the age distributions were generally aligned with expected reproductive staging ^2^. Compared with employed participants, unemployed participants had lower education level, but were otherwise broadly comparable (see Supplementary Table 7 for detailed comparison).

**Table 1.** Demographic characteristics of employed participants by reproductive stage.

|  | Premenopausal<br>(N=255) | Perimenopause<br>(N=485) | Post<br>menopausal<br>(N=83) | Unsure<br>(N=122) | Overall<br>(N=945) |
| --- | --- | --- | --- | --- | --- |
| **Age group** |  |  |  |  |  |
| 35-39 | 64 (25.1%) | 28 (5.8%) | 1 (1.2%) | 27 (22.1%) | 120 (12.7%) |
| 40-44 | 88 (34.5%) | 122 (25.2%) | 3 (3.6%) | 45 (36.9%) | 258 (27.3%) |
| 45-49 | 82 (32.2%) | 249 (51.3%) | 10 (12.0%) | 45 (36.9%) | 386 (40.8%) |
| 50-54 | 18 (7.1%) | 75 (15.5%) | 36 (43.4%) | 5 (4.1%) | 134 (14.2%) |
| 55-59 | 3 (1.2%) | 11 (2.3%) | 33 (39.8%) | 0 (0%) | 47 (5.0%) |
| **Education level** |  |  |  |  |  |
| High school graduate | 61 (23.9%) | 101 (20.8%) | 18 (21.7%) | 31 (25.4%) | 211 (22.3%) |
| Less than high school | 2 (0.8%) | 0 (0%) | 0 (0%) | 2 (1.6%) | 4 (0.4%) |
| Some college or associate degree | 40 (15.7%) | 84 (17.3%) | 16 (19.3%) | 18 (14.8%) | 158 (16.7%) |
| Bachelor's degree | 93 (36.5%) | 174 (35.9%) | 30 (36.1%) | 41 (33.6%) | 338 (35.8%) |
| Graduate or professional degree | 58 (22.7%) | 125 (25.8%) | 19 (22.9%) | 30 (24.6%) | 232 (24.6%) |
| Missing | 1 (0.4%) | 1 (0.2%) | 0 (0%) | 0 (0%) | 2 (0.2%) |
| **Body Mass Index (BMI)** |  |  |  |  |  |
| Normal weight | 100 (39.2%) | 157 (32.4%) | 34 (41.0%) | 44 (36.1%) | 335 (35.4%) |
| Underweight | 5 (2.0%) | 9 (1.9%) | 1 (1.2%) | 5 (4.1%) | 20 (2.1%) |
| Overweight | 63 (24.7%) | 140 (28.9%) | 22 (26.5%) | 35 (28.7%) | 260 (27.5%) |
| Obesity | 84 (32.9%) | 177 (36.5%) | 25 (30.1%) | 38 (31.1%) | 324 (34.3%) |
| Missing | 3 (1.2%) | 2 (0.4%) | 1 (1.2%) | 0 (0%) | 6 (0.6%) |
| **Ethnicity** |  |  |  |  |  |
| White | 164 (64.3%) | 373 (76.9%) | 62 (74.7%) | 86 (70.5%) | 685 (72.5%) |
| Asian | 16 (6.3%) | 17 (3.5%) | 1 (1.2%) | 2 (1.6%) | 36 (3.8%) |
| Black | 46 (18.0%) | 55 (11.3%) | 12 (14.5%) | 15 (12.3%) | 128 (13.5%) |
| Hispanic | 8 (3.1%) | 11 (2.3%) | 3 (3.6%) | 3 (2.5%) | 25 (2.6%) |
| Mixed | 16 (6.3%) | 26 (5.4%) | 3 (3.6%) | 12 (9.8%) | 57 (6.0%) |
| Other | 5 (2.0%) | 3 (0.6%) | 2 (2.4%) | 4 (3.3%) | 14 (1.5%) |

### Labour market participation

Overall, 75.3% of participants in our study sample were in paid employment, broadly comparable with the 72.8% employment-to-population ratio among U.S. women aged 20-64 reported in the 2024 American Community Survey ^39^. Perimenopausal women were equally likely to stay in the workforce compared with those who were premenopausal as labour market participation was broadly comparable across reproductive stages (χ²(3) = 6.67, *p* = .083). Approximately 78.0% of premenopausal and 76.6% of perimenopausal women were in paid employment, compared with 70.3% of postmenopausal women. Contractual hours per week were also similar across reproductive stages: premenopausal, 36.3 (SD = 9.9); perimenopausal, 36.3 (SD = 10.5); postmenopausal, 36.6 (SD = 9.9); and unsure, 34.8 (SD = 10.5), *F*(3) = 0.763, *p* = .52.

### Work and activity impairment by reproductive stage

As shown in Figure 2, work impairment varied significantly by reproductive stage (Kruskal-Wallis χ²(3) = 48.83, *p* < .001).

**Figure 2.**
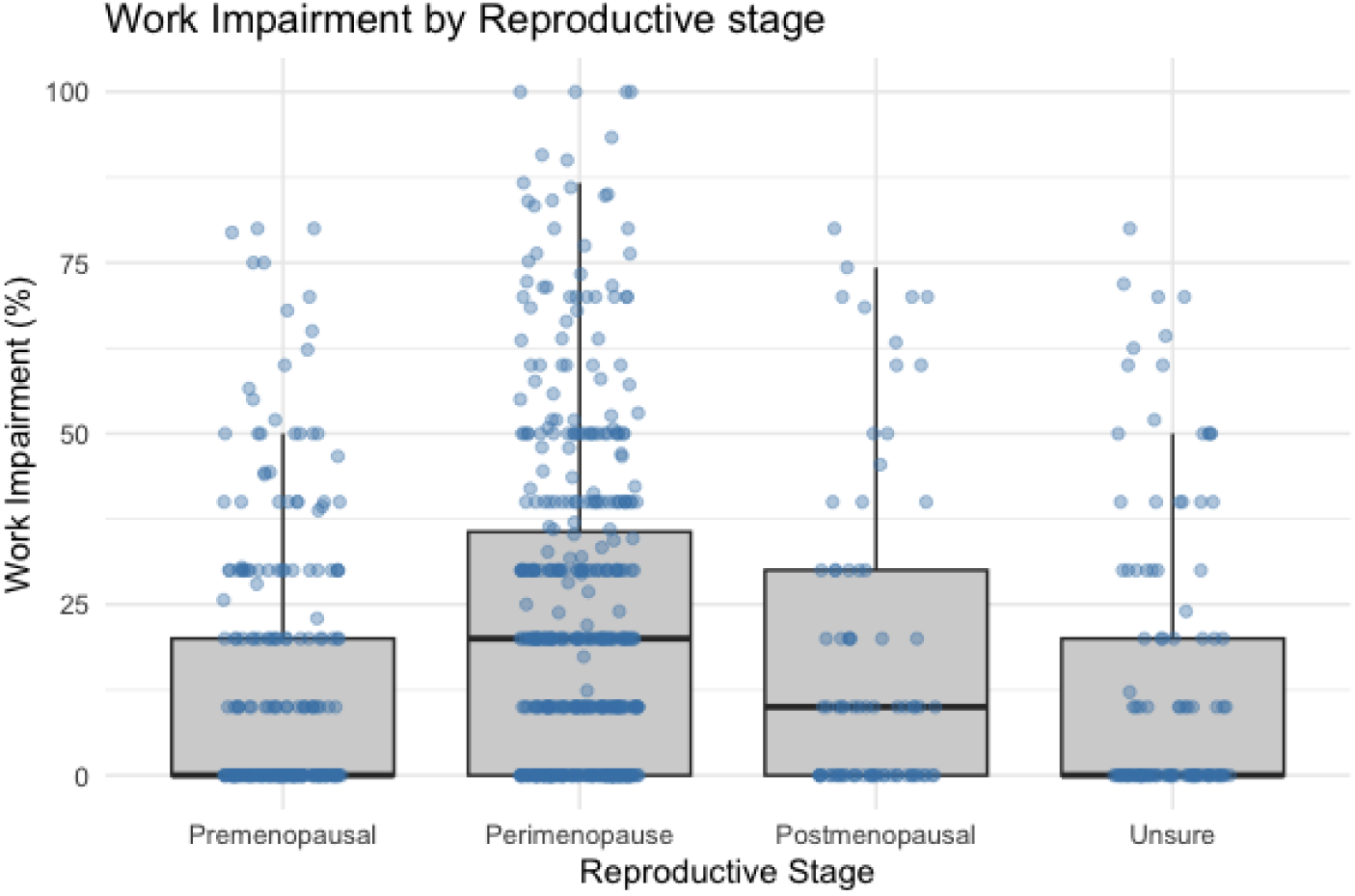
Work impairment by reproductive stage. The distribution of self-reported work impairment (%) as measured by the Work Productivity and Activity Impairment (WPAI) questionnaire, by reproductive stage.

Women in perimenopause consistently reported the highest levels of work and activity impairment across reproductive stages (Table 2). Mean work impairment reached 22.5% in perimenopausal women, compared with 12.7% in premenopausal women,16.9% in postmenopausal women, and 13.7% among women who were unsure of their reproductive stage. The observed difference between premenopausal and perimenopausal women was 9.8 percentage points, and the difference remained substantial at 7.92 percentage points (95% CI: 4.3% – 11.5%) after adjusting for age, BMI, education, and ethnicity (see Supplementary Table 2a and Table 3). A similar pattern was observed for activity impairment, which was also highest among perimenopausal women at 23.5% (Kruskal-Wallis χ²(3) = 46.09, *p* < .001).

**Table 2.** Work and activity impairment by self-reported reproductive stage.

|  | Premenopausal<br>(N=255) | Perimenopause<br>(N=485) | Postmenopausal<br>(N=83) | Unsure<br>(N=122) | Overall<br>(N=945) |
| --- | --- | --- | --- | --- | --- |
| Absenteeism (%) |  |  |  |  |  |
| Mean (SD) | 1.90 (7.70) | 2.94 (9.29) | 1.66 (7.03) | 1.20 (5.77) | 2.32 (8.32) |
| Median [Min, Max] | 0 [0, 70.6] | 0 [0, 66.7] | 0 [0, 40.0] | 0 [0, 43.8] | 0 [0, 70.6] |
| Missing | 5 (2.0%) | 14 (2.9%) | 3 (3.6%) | 6 (4.9%) | 28 (3.0%) |
| **Presenteeism (%)** |  |  |  |  |  |
| Mean (SD) | 11.5 (17.3) | 20.9 (22.2) | 16.0 (21.7) | 12.9 (19.7) | 16.9 (21.0) |
| Median [Min, Max] | 0 [0, 80.0] | 20.0 [0, 100] | 10.0 [0, 80.0] | 0 [0, 80.0] | 10.0 [0, 100] |
| **Work impairment (%)** |  |  |  |  |  |
| Mean (SD) | 12.7 (19.3) | 22.5 (24.0) | 16.9 (22.9) | 13.7 (21.0) | 18.2 (22.7) |
| Median [Min, Max] | 0 [0, 80.0] | 20.0 [0, 100] | 10.0 [0, 80.0] | 0 [0, 80.0] | 10.0 [0, 100] |
| Missing | 5 (2.0%) | 14 (2.9%) | 3 (3.6%) | 6 (4.9%) | 28 (3.0%) |
| **Activity impairment (%)** |  |  |  |  |  |
| Mean (SD) | 13.1 (18.5) | 23.5 (23.9) | 20.7 (24.6) | 16.1 (22.1) | 19.5 (22.8) |
| Median [Min, Max] | 0 [0, 80.0] | 20.0 [0, 100] | 10.0 [0, 90.0] | 0 [0, 80.0] | 10.0 [0, 100] |

**Note.** Values are Work Productivity and Activity Impairment (WPAI) scores expressed as percentages, ranging from 0 to 100%, with higher scores indicating greater impairment.

**Table 3.**
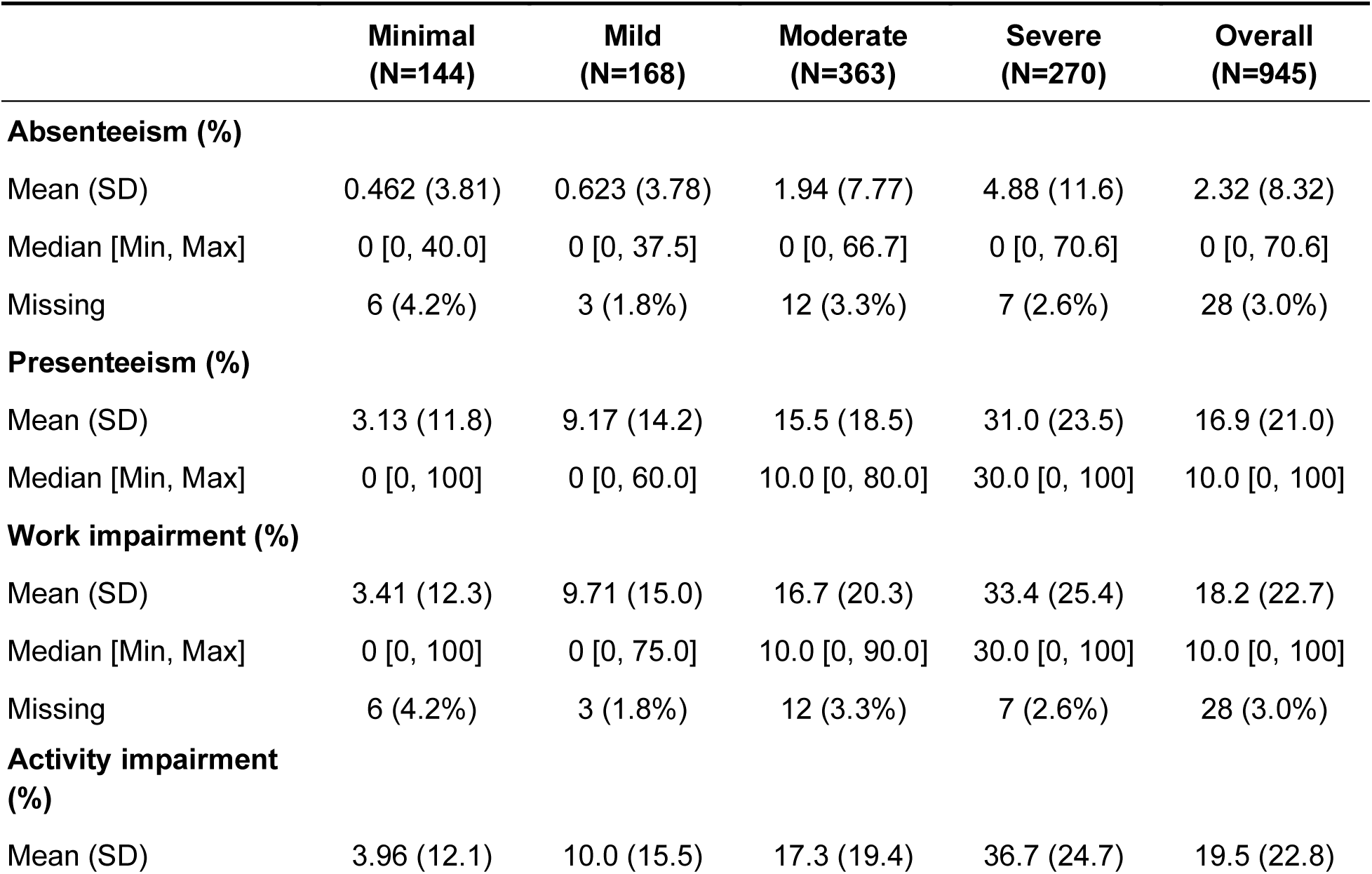
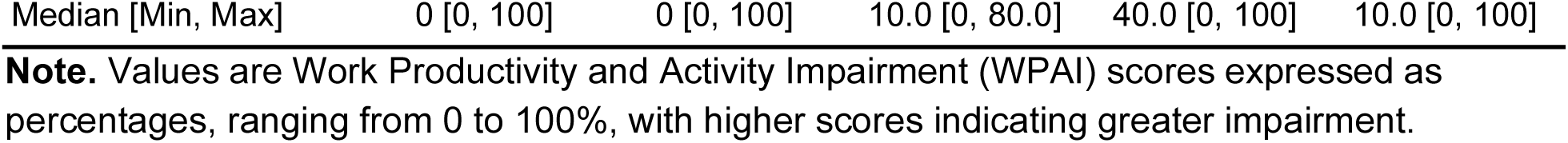
Work and activity impairment by symptom severity (as measured by the Menopause Rating Scale)

### Work and activity impairment by symptom severity

Symptom severity was also associated with workplace impairment. As shown in Figure 3 and Table 3, a clear and statistically significant gradient was observed between work impairment and symptom severity (Kruskal–Wallis χ²(3) = 226.31, *p* < .001). This pattern remained after adjustment for age group, BMI, education, and ethnicity (Supplementary Table 2b).

**Figure 3.**
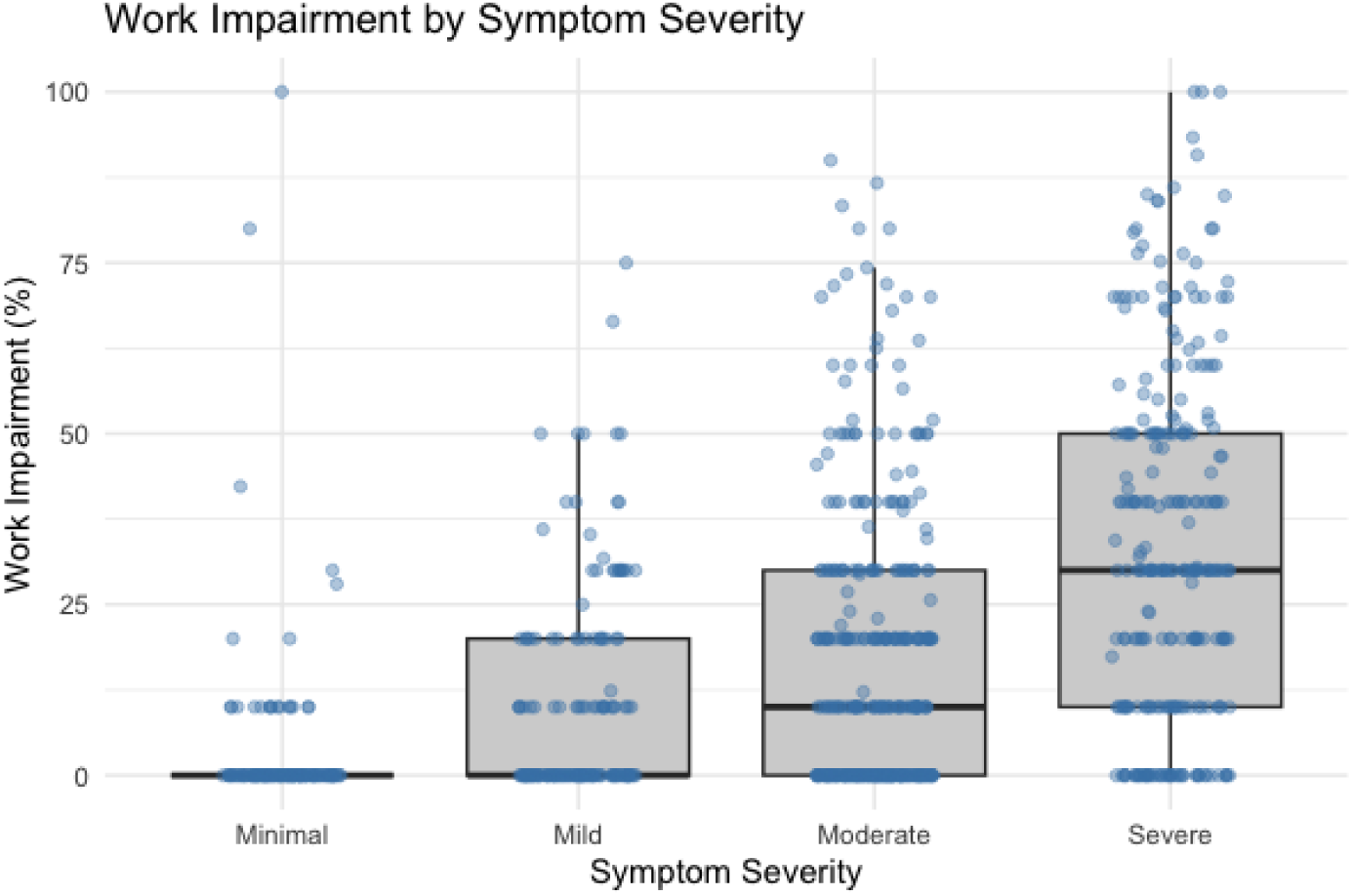
Work impairment by symptom severity. The distribution of self-reported work impairment (%) as measured by the Work Productivity and Activity Impairment questionnaire, by symptom severity as measured by the Menopause Rating Scale.

The mean work impairment increased from 3.4% among women with minimal symptoms to 9.7% with mild symptoms, 16.7% with moderate symptoms, and 33.4% among those with severe symptoms. Notably, impairment was already substantial among women with moderate symptom burden, emerging well before the most severe symptom levels. Compared with women with minimal symptoms, the observed differences were 6.3, 13.3, and 30.0 percentage points for mild, moderate, and severe symptoms, respectively, and the differences remained at similar levels after adjusting for demographics (Supplementary Table 2b).

Similarly, activity impairment varied significantly by symptom severity (Kruskal–Wallis χ²(3) = 273.92, p < 0.001), increasing from 4.0% in the minimal symptom group to 36.7% among those with severe symptoms.

When decomposing work impairment, the majority was driven by presenteeism rather than absenteeism. Across symptom severity groups, absenteeism remained low, increasing modestly from 0.46% among women with minimal symptoms to 4.88% among those with severe symptoms, whereas presenteeism rose markedly from 3.13% to 31.0% (Table 3).

### Symptom domains associated with work impairment

Not all symptom domains had an equal impact on work impairment (Table 4). Somatic symptoms were the strongest predictor of work impairment (β = 2.44, 95% CI: 1.81–3.07, *p* < .001), followed by psychological symptoms (β = 1.28, 95% CI: 0.80–1.76, *p* < .001), adjusted for age, BMI, education, and ethnicity. In contrast, the effects of urogenital symptoms appeared non-significant (*p* = .27). The results were consistent with associations at individual symptom level (see Supplementary Table 1).

**Table 4.** Multivariable linear regression examining the effects of the Menopause Rating Scale (MRS) symptom domains on work impairment.

| Predictors | Work Impairment |  |  |
| --- | --- | --- | --- |
|  | Estimates | 95% CI | p value |
| (Intercept) | -7.47** | -12.72 – -2.21 | **0.005** |
| **MRS domain** |  |  |  |
| Psychological score | 1.28*** | 0.80 – 1.76 | **<0.001** |
| Somatic score | 2.44*** | 1.81 – 3.07 | **<0.001** |
| Urogenital score | 0.36 | -0.28 – 1.00 | 0.273 |
| **Age group (ref = 35-39)** |  |  |  |
| 40-44 | 3.77 | -0.61 – 8.15 | 0.092 |
| 45-49 | 6.08** | 1.91 – 10.25 | **0.004** |
| 50-54 | 5.82* | 0.71 – 10.92 | **0.026** |
| 55-59 | 0.23 | -6.64 – 7.11 | 0.947 |
| **Education (ref = High school graduate)** |  |  |  |
| Less than high school | 0.59 | -18.93 – 20.11 | 0.953 |
| Some college or associate degree | 3.09 | -1.10 – 7.27 | 0.148 |
| Bachelor's degree | 4.08* | 0.56 – 7.59 | **0.023** |
| Graduate or professional degree | 1.94 | -1.88 – 5.75 | 0.319 |
| **Body Mass Index (BMI)** |  |  |  |
| **(ref = Normal weight)** |  |  |  |
| Underweight | -1.84 | -10.98 – 7.29 | 0.692 |
| Overweight | 0.75 | -2.52 – 4.02 | 0.654 |
| Obese | -0.37 | -3.52 – 2.78 | 0.817 |
| **Ethnicity (ref = White)** |  |  |  |
| Asian | -6.35 | -13.04 – 0.34 | 0.063 |
| Black | 2.52 | -1.36 – 6.40 | 0.203 |
| Hispanic | -3.61 | -11.48 – 4.26 | 0.368 |
| Mixed | 6.82* | 1.44 – 12.19 | **0.013** |

| Other | 0 | -10.42 – 10.43 | 0.999 |
| --- | --- | --- | --- |
| Observations | 910 |  |  |
| R<sup>2</sup> / R<sup>2</sup> adjusted | 0.270 / 0.251 |  |  |

\* p < .05; \*\* p < .01; \*\*\* p < .001.
**Note.** The outcome was overall work impairment measured using the WPAI and expressed as a percentage (0–100%). $\beta$ coefficients represent the adjusted difference in work impairment, in percentage points, associated with a one-point increase in the respective MRS domain score.

### Economic impact of perimenopause

The observed work impairment associated with perimenopause and symptom burden translated into substantial economic losses at both the individual and societal levels.

At the individual level, compared with premenopausal women, for a typical perimenopausal woman earning the median salary, the estimated annual productivity loss reached approximately $6,061 per person (equivalent to 9.8% of annual salary), assuming median weekly earnings for U.S. women aged 45-54 at $1,193 in 2024 ^16^.

The productivity loss increased markedly with symptom severity. Compared with women with minimal or no symptoms, the annual productivity loss of those with mild symptoms was estimated at $3,908, rising to $8,257 among those with moderate symptoms and $18,586 among those with severe symptoms.

When demographic factors were taken into account, the productivity loss associated with perimenopause was reduced to approximately $4,900.84 per person. In contrast, the productivity loss estimates by symptom severity changed only modestly after adjusting for demographic factors (Supplementary Table 5).

Applying the individual-level estimates to the U.S. female workforce, the societal productivity loss was estimated to be approximately $56.7 billion per year (see Supplementary Table 6 for extrapolation process), or $45.9 billion if adjusting for demographic factors.

In addition to work and activity impairment outcomes, we also examined health-related quality of life as measured by EuroQoL EQ-5D-5L using U.S. specific tariffs ^32^. Mean utility was 0.862 (SD = 0.165) among premenopausal women and 0.805 (SD = 0.185) among perimenopausal women. Utility decreased progressively with symptom severity, from 0.966 (SD = 0.055) among women with minimal symptoms to 0.712 (SD = 0.214) among those with severe symptoms. Regression-adjusted estimates are provided in Supplementary Tables 4 to support future economic evaluations.

## Discussion

### Principal findings

This study is the first, to our knowledge, to quantify the workplace productivity impact and economic burden of perimenopause among U.S. women. Although labour market participation was similar between perimenopausal and premenopausal women (76.6% vs. 78.0%), perimenopausal women reported significantly higher work impairment (22.5%) than women of other reproductive stages (premenopausal: 12.7%; postmenopausal: 16.9%; unsure: 13.7%). Work impairment was largely driven by presenteeism rather than absenteeism, and it increased with symptom severity level, rising from 3.4% among women with minimal symptoms to 33.4% among those with severe symptoms.

Somatic and psychological symptoms were most strongly associated with productivity loss, whereas the effects of urogenital symptoms appeared non-significant, after adjusting for demographic characteristics.

These productivity losses translate into a substantial economic burden at both individual and societal level. For a typical perimenopausal woman in the U.S. labour force earning the median salary, the estimated productivity loss amounted to approximately $6,061 per year (9.8% of annual salary). Extrapolated to the U.S. workforce, this corresponds to an annual productivity loss of $56.7 billion.

### Workplace and economic implications of perimenopause

Perimenopause may represent a hidden economic burden that is distinct from those captured in previous studies. Unlike the “menopause penalty”, which has largely been examined through the lens of changes in labour market participation, earnings, and retirement timing decisions ^12^, our findings suggest productivity losses associated with perimenopause arise mainly from increased work impairment among women who remain in the labour force. A similar contrast can be drawn with the “motherhood penalty”, which is often linked to career breaks, reduced working hours, or barriers to return to the labour market that are associated with childbirth and caregiving responsibilities ^40–42^. In comparison, our findings indicate that the economic burden associated with perimenopause appears to be embedded within continued employment, making it less visible to conventional labour market statistics, and thus less likely to attract institutional recognition or policy attention. The lower visibility is particularly notable, given that the burden (9.8% of annual earnings) is comparable to the motherhood wage penalty, which was estimated to be around 7% annual earnings per child among U.S. women ^43^.

The fact that perimenopause productivity loss is primarily presenteeism-driven may also help explain why our estimates of productivity losses associated with perimenopause were much higher than previous estimates for menopausal symptoms. According to Faubion and colleagues ^18^, the estimated annual workplace productivity loss due to menopausal symptoms, based solely on absenteeism, stood at $1.8 billion, corresponding to approximately 1.2% of annual workdays missed. In contrast, our estimate was based on overall work impairment of 9.8%, capturing both absenteeism and presenteeism. Our estimate also reflects a larger potentially affected population. As perimenopausal symptoms may begin in the mid-30s ^22^, the affected population spans a wider working-age range than the menopausal workforce. In addition, we included women in both full-time and part-time jobs, whereas Faubion and colleagues restricted to menopausal population in full-time jobs.

The productivity loss of perimenopause may also be overlooked for lack of clear and recognisable starting points, compared with other major reproductive transitions ^1,44^. Motherhood is a highly visible life event, while menopause, although more private, has a definitive retrospective confirmation point after 12 months without menstruation. The visibility and certainty that accompany these transitions make their labour market effects easier to observe and measure. Perimenopause, by contrast, comes with much higher uncertainty, particularly at the early transition period before women have a recognised label for what is happening to them ^10^. This uncertainty may be further compounded by stigma surrounding disclosure of symptoms, potentially discouraging women from seeking support at workplace ^45^. The productivity loss of perimenopause may therefore accumulate due to delays in care and support seeking. In this sense, perimenopause may represent a rare form of labour-market burden that is simultaneously common, costly, and difficult to observe: normalised as stress or ageing, experienced privately, and only partially captured by conventional labour market statistics.

Beyond direct productivity losses, perimenopause may also incur opportunity costs. As this life stage commonly coincides with women’s peak-earning years in their careers, persistent symptoms could reduce women’s workplace identity, confidence, aspirations, professional development, and career progression ^46^. While these downstream effects were not measured in our study, they suggest that the full economic significance of perimenopause may be even higher than our estimate.

### Symptom-driven work impairment and implications for workplace support

Our findings also suggest that perimenopause-related productivity loss is more than a simple story of “higher symptom severity, less work productivity” – some symptom domains appeared more consequential than others. One possible explanation is that symptoms only affect productivity when they collide with the specific demands of a person’s work ^47,48^.

The stronger associations we observed for somatic and psychological symptoms suggest these domains may be most disruptive to core workplace functioning. Somatic symptoms such as musculoskeletal pain may reduce physical endurance, vigilance, and attention ^49^, whereas psychological symptoms such as irritability or anxiety might affect concentration, emotional regulation and executive decision-making ^50^. Those symptom-specific mechanisms may also explain why urogenital symptoms (e.g. vaginal dryness) were not significantly associated with work impairment, although they may still affect women’s quality of life.

These findings have important implications for workplace support. The Job Demands-Resources theory posits that the negative impacts of demanding jobs can be buffered when workers are offered more resources such as autonomy, flexibility, and leadership support ^45,46^. Applied to perimenopause, our findings imply that productivity loss is not inevitable when symptoms arise. Perimenopausal women are not ***inherently*** less productive, and the symptom-driven productivity loss might well be prevented or minimised if they are well supported. While existing workplace recommendations have largely focused on increasing awareness, reducing stigma, and improving access to flexible working or occupational health support ^49^, our findings suggest that workplace support might be more effective when they match women’s symptom profiles and job demands. For example, flexible working may help women experiencing physical and mental exhaustion or sleep disruption in cognitively demanding roles, whereas temperature control, flexibility in dress code, or additional work breaks might help women experiencing vasomotor symptoms in physically demanding or poorly ventilated work environments ^51,52^.

Access to workplace support, however, may be unevenly distributed. Frontline workers or those in lower-paid jobs may have less access to flexible or remote working arrangements and other workplace resources that could mitigate symptom-related impairment ^53,54^. Workplace support strategies should therefore consider not only effectiveness but also equitable access.

Last but not least, as perimenopausal symptom profiles may fluctuate and evolve over time ^21,55^, the type and intensity of workplace support women require may need to be dynamic and adaptable. Manager education, occupational health involvement, and customizable workplace-based interventions may facilitate adjustments as women’s needs change throughout the transition ^51,52,56^. Digital symptom tracking and monitoring tools ^57,58^ may further complement workplace and clinical support by helping women identify symptom patterns, monitor trends, and align support with their evolving needs.

### Strengths, limitations and future directions

This study has several strengths. It addresses an important yet under-researched challenge that impacts midlife women at the workplace. By quantifying the workplace productivity loss of perimenopause and identifying the main symptom drivers, it provides an initial evidence base for policy responses and design of workplace support. Compared with previous studies ^19,51^, we used validated instruments to measure both symptom burden and work impairment, enabling comparison with productivity losses associated with other reproductive life stages and health conditions.

This study also has several limitations. First, the cross-sectional design limits causal inferences as it only captures a snapshot of women’s symptom burden and work impairment, which might not be representative of perimenopause. However, the wide range of age, symptom burden, and work impairment observed in our sample may partially mitigate this limitation. Second, the reproductive stage was self-reported and may be misclassified, although previous studies using this methodology ^23^ suggest reasonable validity of this approach and observed symptom-severity patterns (Figure 1) were broadly consistent with expected reproductive-stage differences. Third, although Prolific employs participant verification and data-quality safeguards, including identity verification and controls to detect duplicate accounts, some eligibility criteria relied on self-report and intentional misrepresentation cannot be entirely excluded. In addition, information on participants’ occupation sectors, job characteristics, and workplace environments or facilities was not collected, which limited our ability to assess how these contextual factors may have influenced symptom-related work impairment. Finally, our economic estimates should be interpreted as applying to a typical perimenopausal woman in our study population. Although the proportion of participants in paid employment (75.3%) was broadly comparable with the 72.8% employment-to-population ratio among U.S. women aged 20-64 ^39^, the sample was not nationally representative, which might limit the generalisability to the wider U.S. workforce. Population-level productivity-loss estimates should consequently be interpreted as model-based approximations.

Future research should move beyond cross-sectional snapshots. Longitudinal cohort studies that track women’s symptom burden and work impairment across reproductive stages could identify periods of heightened vulnerability and examine whether changes in symptom burden precede changes in work impairment, strengthening causal inference. Another direction is to investigate downstream consequences not captured in this study, including effects of perimenopausal symptoms on career aspirations, earnings trajectories, and other opportunity costs. Finally, impact evaluations of tailored workplace support are also needed to determine how much of the observed productivity loss may be preventable and which approaches (e.g. flexible working, adjustable workload, occupational health support) are most effective in reducing symptom-related productivity loss.

## Conclusion

Perimenopause may represent a substantial yet overlooked workplace productivity challenge for midlife women in the United States. The hidden productivity toll of perimenopause appears not because women leave the workforce, but because they continue working while managing symptoms. Work impairment increases sharply with symptom severity and is driven primarily by somatic and psychological symptoms, translating into significant productivity losses at both individual and societal levels. Recognising and quantifying this productivity toll may be the first step towards offering more dedicated support for perimenopausal women at the workplace, which requires coordinated efforts from employers, policymakers, insurers, healthcare providers, and digital health innovators.

## Abbreviations

BMI: Body Mass Index
BLS: Bureau of Labor Statistics
CI: Confidence Interval
EQ-5D-5L: EuroQol five-dimensional, five-level questionnaire
HRQoL: Health-related quality of life
IRB: Institutional Review Board
MRS: Menopause Rating Scale
SD: Standard Deviation
WPAI: Work Productivity and Activity Impairment questionnaire

## Acknowledgements

We would like to thank all the individuals who participated in our study. We would also like to acknowledge the valuable suggestions and feedback received from attendees of the 2025 Annual Meeting of The Menopause Society, where an earlier version of this work was presented as an abstract.

## Author contributions

Y.X. conceived the study, developed the study protocol, designed the survey, conducted the statistical analyses, and drafted the manuscript. C.P. co-developed the study protocol, led the ethics application and survey implementation, and contributed to the literature review. Y.H-M. refined the study protocol, supported the ethics application, and contributed to the literature review. A.C.C. contributed to the design and implementation of the survey. L.Z. refined the study conceptualisation and supervised the study design and survey implementation. J.P.J. supervised the study methodology and statistical analyses and critically revised the manuscript.

All authors reviewed and approved the final manuscript.

## Declarations and Statements

### Data availability

The deidentified data that support the findings of this study are available upon reasonable request and with the permission of Flo Health UK Limited.

### Code availability

Data was cleaned, aggregated, and analysed using SQL and R version 4.4.1. The R code is available upon reasonable request from the corresponding author and with the permission of Flo Health UK Limited.

## Supplementary materials

**Supplementary Table 1.**
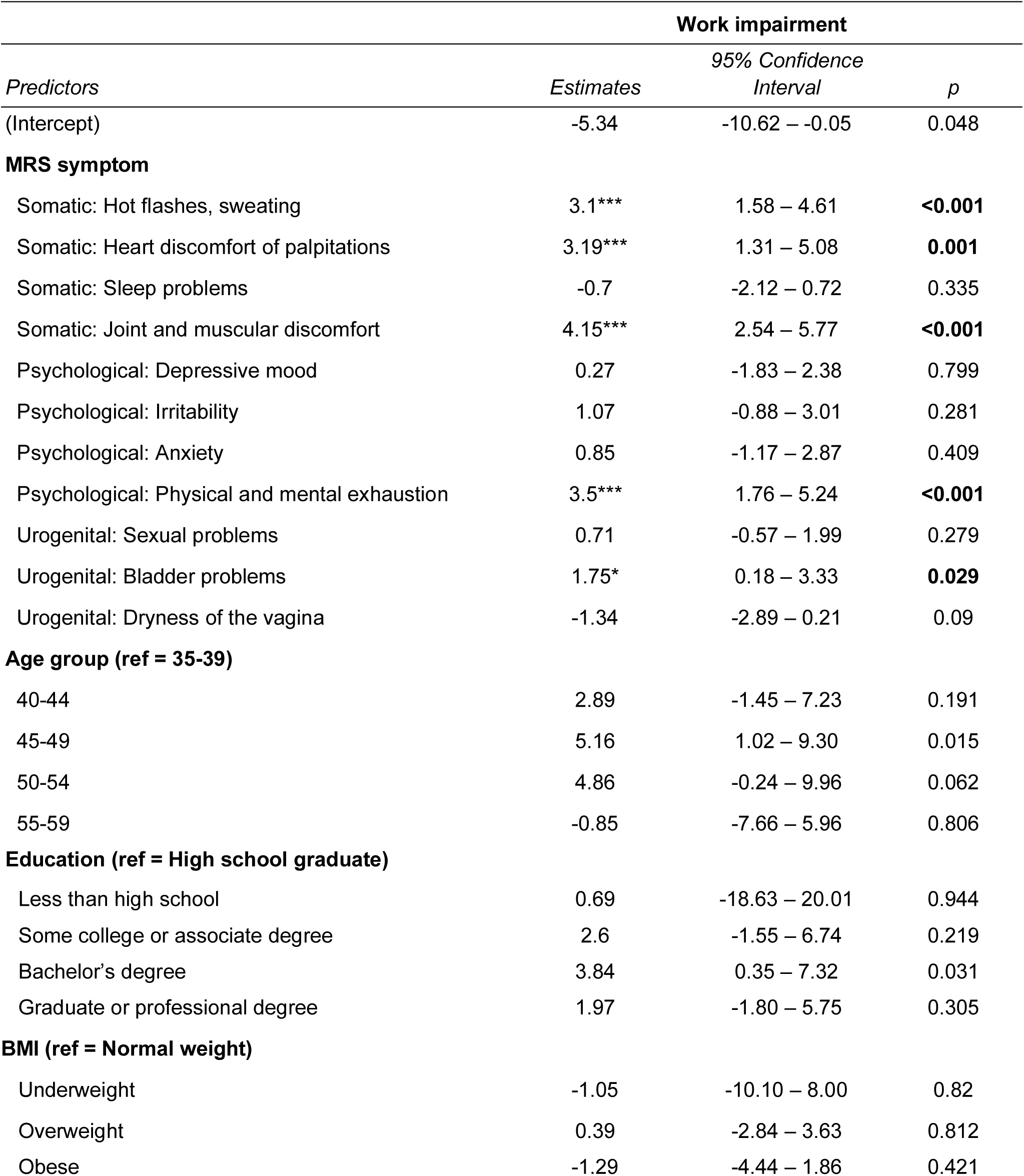
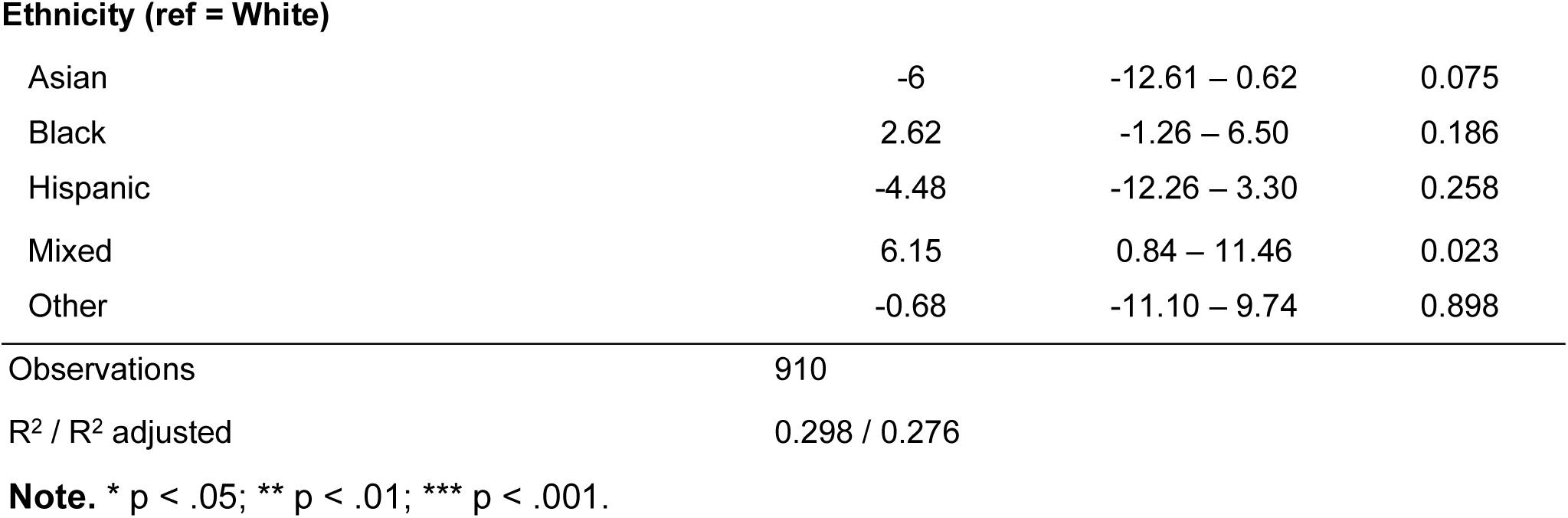
Multivariable linear regression examining the effects of individual MRS symptom item on work impairment.

| *Predictors* | **Work impairment** |  |  |
| --- | --- | --- | --- |
|  | *Estimates* | *95% Confidence Interval* | *p* |
| (Intercept) | -5.34 | -10.62 – -0.05 | 0.048 |
| **MRS symptom** |  |  |  |
| Somatic: Hot flashes, sweating | 3.1*** | 1.58 – 4.61 | <0.001 |
| Somatic: Heart discomfort or palpitations | 3.19*** | 1.31 – 5.08 | 0.001 |
| Somatic: Sleep problems | -0.7 | -2.12 – 0.72 | 0.335 |
| Somatic: Joint and muscular discomfort | 4.15*** | 2.54 – 5.77 | <0.001 |
| Psychological: Depressive mood | 0.27 | -1.83 – 2.38 | 0.799 |
| Psychological: Irritability | 1.07 | -0.88 – 3.01 | 0.281 |
| Psychological: Anxiety | 0.85 | -1.17 – 2.87 | 0.409 |
| Psychological: Physical and mental exhaustion | 3.5*** | 1.76 – 5.24 | <0.001 |
| Urogenital: Sexual problems | 0.71 | -0.57 – 1.99 | 0.279 |
| Urogenital: Bladder problems | 1.75* | 0.18 – 3.33 | 0.029 |
| Urogenital: Dryness of the vagina | -1.34 | -2.89 – 0.21 | 0.09 |
| **Age group (ref = 35-39)** |  |  |  |
| 40-44 | 2.89 | -1.45 – 7.23 | 0.191 |
| 45-49 | 5.16 | 1.02 – 9.30 | 0.015 |
| 50-54 | 4.86 | -0.24 – 9.96 | 0.062 |
| 55-59 | -0.85 | -7.66 – 5.96 | 0.806 |
| **Education (ref = High school graduate)** |  |  |  |
| Less than high school | 0.69 | -18.63 – 20.01 | 0.944 |
| Some college or associate degree | 2.6 | -1.55 – 6.74 | 0.219 |
| Bachelor's degree | 3.84 | 0.35 – 7.32 | 0.031 |
| Graduate or professional degree | 1.97 | -1.80 – 5.75 | 0.305 |
| **BMI (ref = Normal weight)** |  |  |  |
| Underweight | -1.05 | -10.10 – 8.00 | 0.82 |
| Overweight | 0.39 | -2.84 – 3.63 | 0.812 |
| Obese | -1.29 | -4.44 – 1.86 | 0.421 |
| **Ethnicity (ref = White)** |  |  |  |
| Asian | -6 | -12.61 – 0.62 | 0.075 |
| Black | 2.62 | -1.26 – 6.50 | 0.186 |
| Hispanic | -4.48 | -12.26 – 3.30 | 0.258 |
| Mixed | 6.15 | 0.84 – 11.46 | 0.023 |
| Other | -0.68 | -11.10 – 9.74 | 0.898 |
| Observations | 910 |  |  |
| R<sup>2</sup> / R<sup>2</sup> adjusted | 0.298 / 0.276 |  |  |

**Note.** \* p < .05; \*\* p < .01; \*\*\* p < .001.

**Supplementary Table 2a.**
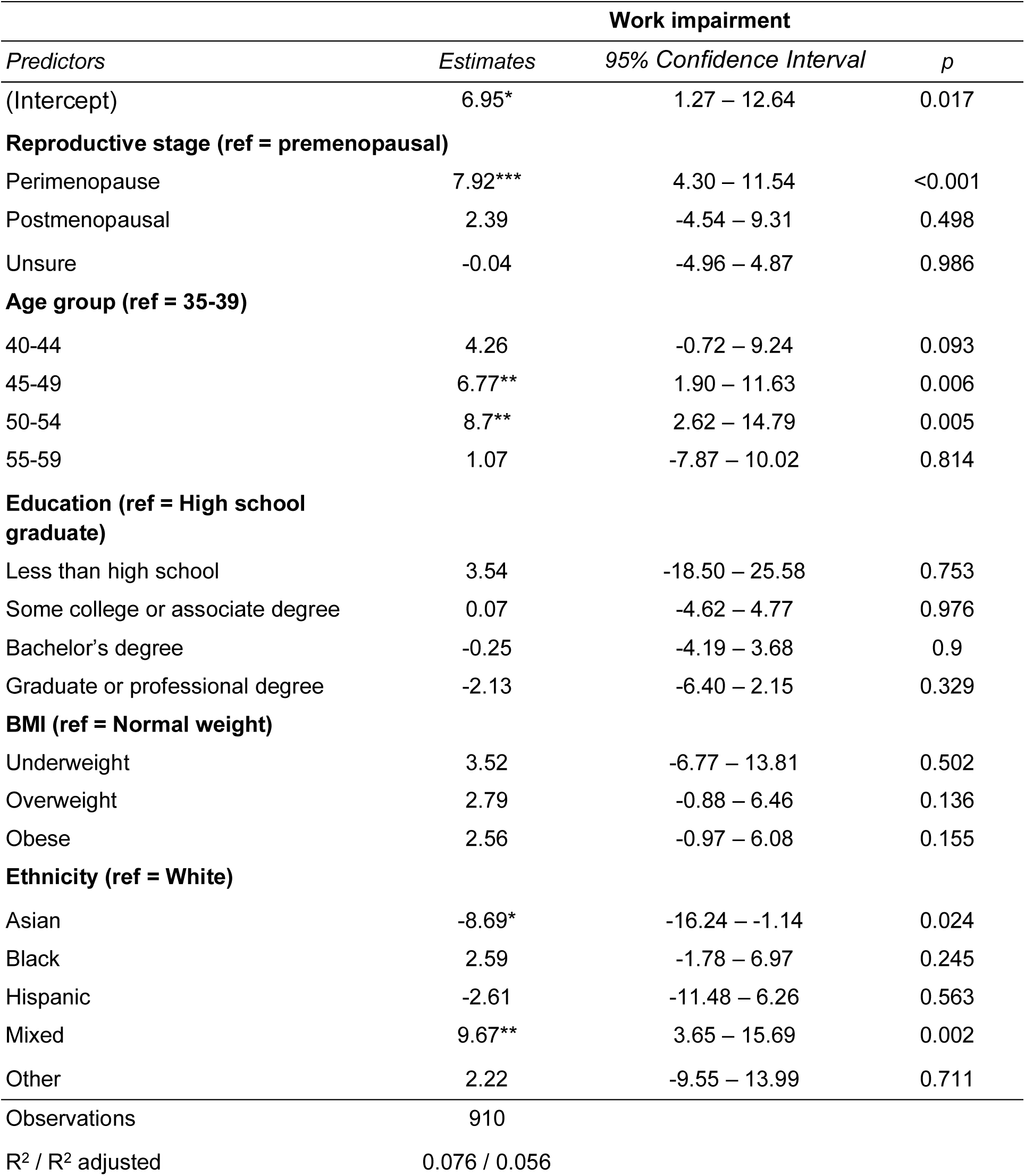
Multivariable linear regression examining how work impairment varies by reproductive stage.

**Supplementary Table 2b.** Multivariable linear regression examining how work impairment varies by symptom burden as measured by Menopause Rating Scale (MRS)

| *Predictors* | **Work impairment** |  |  |
| --- | --- | --- | --- |
|  | *Estimates* | *CI* | *p* |
| (Intercept) | -2.96 | -8.56 – 2.64 | 0.3 |
| **Symptom burden (ref = Minimal)** |  |  |  |
| Mild | 5.33 * | 0.73 – 9.92 | 0.023 |
| Moderate | 12.29 *** | 8.27 – 16.30 | <0.001 |
| Severe | 28.90 *** | 24.67 – 33.13 | <0.001 |
| **Age group (ref = 35-39)** |  |  |  |
| 40-44 | 3.7 | -0.74 – 8.14 | 0.102 |
| 45-49 | 6.53 ** | 2.29 – 10.76 | 0.003 |
| 50-54 | 6.66 * | 1.58 – 11.74 | 0.01 |
| 55-59 | 0.76 | -6.09 – 7.61 | 0.827 |
| **Education (ref = High school graduate)** |  |  |  |
| Less than high school | -0.96 | -20.79 – 18.87 | 0.924 |
| Some college or associate degree | 2.27 | -1.97 – 6.51 | 0.294 |
| Bachelor's degree | 3.17 | -0.38 – 6.73 | 0.08 |
| Graduate or professional degree | 0.67 | -3.19 – 4.53 | 0.735 |
| **BMI (ref = Normal weight)** |  |  |  |
| Underweight | -0.12 | -9.40 – 9.17 | 0.98 |
| Overweight | 1.33 | -1.98 – 4.65 | 0.43 |
| Obese | 0.05 | -3.15 – 3.25 | 0.976 |
| **Ethnicity (ref = White)** |  |  |  |
| Asian | -6.48 | -13.27 – 0.31 | 0.061 |
| Black | 2.41 | -1.53 – 6.35 | 0.231 |
| Hispanic | -3.45 | -11.44 – 4.54 | 0.397 |
| Mixed | 7.23 ** | 1.81 – 12.66 | 0.009 |
| Other | 0.46 | -10.13 – 11.05 | 0.932 |
| Observations | 910 |  |  |
| R<sup>2</sup> / R<sup>2</sup> adjusted | 0.249 / 0.233 |  |  |

**Note.** Work impairment was measured using the Work Productivity and Activity Impairment (WPAI) questionnaire and is expressed as a percentage (0–100%), with higher values indicating greater impairment. Regression estimates represent adjusted differences in work impairment in percentage points relative to the stated reference category, holding other variables in the model constant. \* $p < .05$ ; \*\* $p < .01$ ; \*\*\* $p < .001$ .

**Supplementary Table 3.** Work impairment by reproductive stage and symptom severity (observed and adjusted)

|  | n | Observed mean (% , SD) | Adjusted mean (% , 95% CI) |
| --- | --- | --- | --- |
| **Reproductive stage** |  |  |  |
| Premenopausal | 255 | 12.73 (19.34) | 14.10 (7.73–20.50) |
| Perimenopausal | 485 | 22.50 (23.95) | 22.00 (15.71–28.40) |
| Postmenopausal | 83 | 16.89 (22.85) | 16.50 (8.57–24.40) |
| Unsure | 122 | 13.68 (21.00) | 14.10 (7.21–20.90) |
| **Symptom severity** |  |  |  |
| Minimal | 144 | 3.41 (12.25) | 1.94 (0–8.06) |
| Mild | 168 | 9.71 (14.98) | 7.27 (1.21–13.33) |
| Moderate | 363 | 16.72 (20.25) | 14.23 (8.66–19.80) |
| Severe | 270 | 33.37 (25.41) | 30.84 (25.19–36.49) |

**Note.** Adjusted means were estimated from multivariable linear regression models controlling for age group, BMI, education, and ethnicity. Values represent marginal means with 95% confidence intervals.

**Supplementary Table 4.**
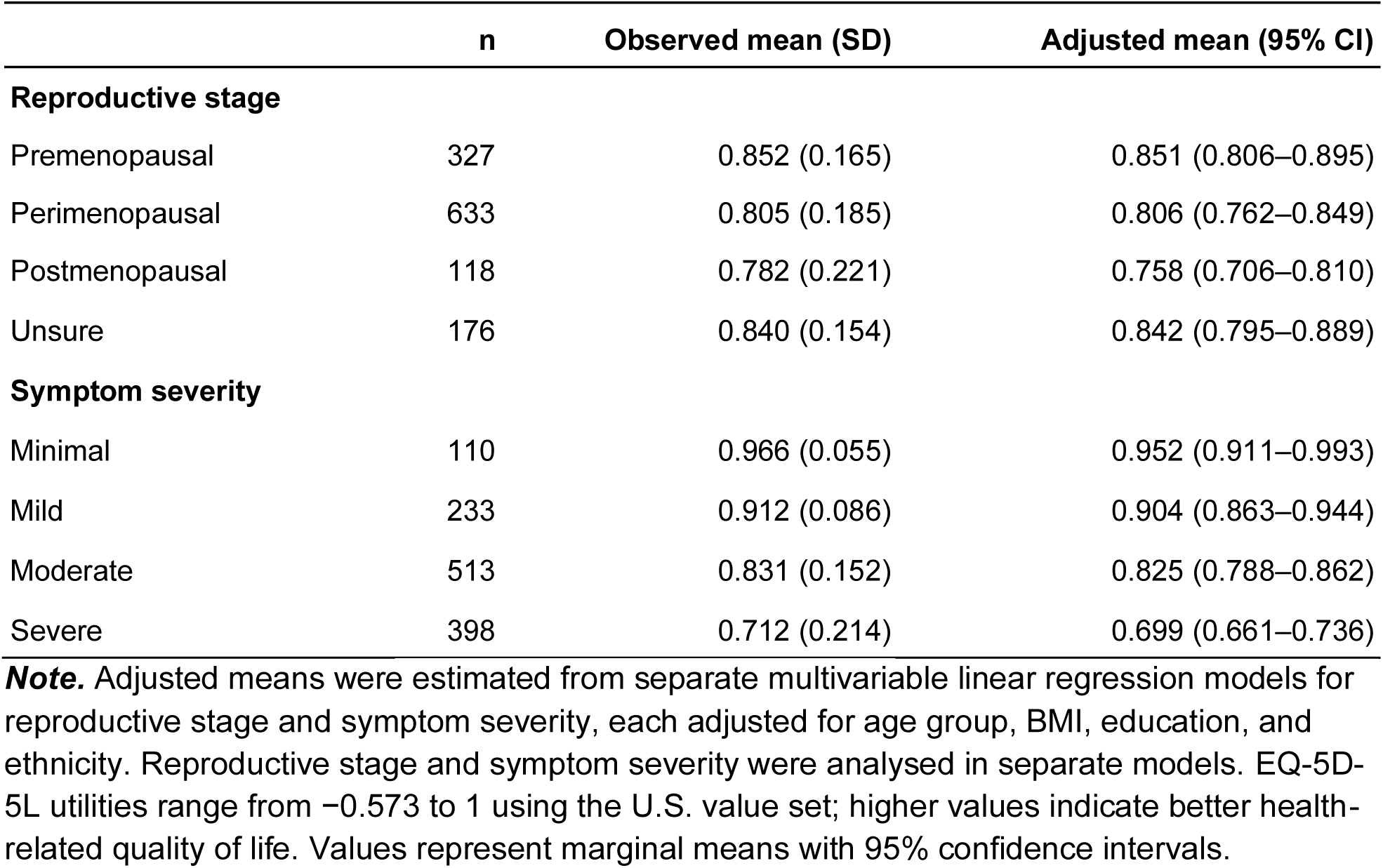
EQ-5D-5L utility by reproductive stage and symptom severity (observed and adjusted)

|  | n | Observed mean (SD) | Adjusted mean (95% CI) |
| --- | --- | --- | --- |
| **Reproductive stage** |  |  |  |
| Premenopausal | 327 | 0.852 (0.165) | 0.851 (0.806–0.895) |
| Perimenopausal | 633 | 0.805 (0.185) | 0.806 (0.762–0.849) |
| Postmenopausal | 118 | 0.782 (0.221) | 0.758 (0.706–0.810) |
| Unsure | 176 | 0.840 (0.154) | 0.842 (0.795–0.889) |
| **Symptom severity** |  |  |  |
| Minimal | 110 | 0.966 (0.055) | 0.952 (0.911–0.993) |
| Mild | 233 | 0.912 (0.086) | 0.904 (0.863–0.944) |
| Moderate | 513 | 0.831 (0.152) | 0.825 (0.788–0.862) |
| Severe | 398 | 0.712 (0.214) | 0.699 (0.661–0.736) |

**Note.** Adjusted means were estimated from separate multivariable linear regression models for reproductive stage and symptom severity, each adjusted for age group, BMI, education, and ethnicity. Reproductive stage and symptom severity were analysed in separate models. EQ-5D-5L utilities range from –0.573 to 1 using the U.S. value set; higher values indicate better health-related quality of life. Values represent marginal means with 95% confidence intervals.

**Supplementary Table 5.**
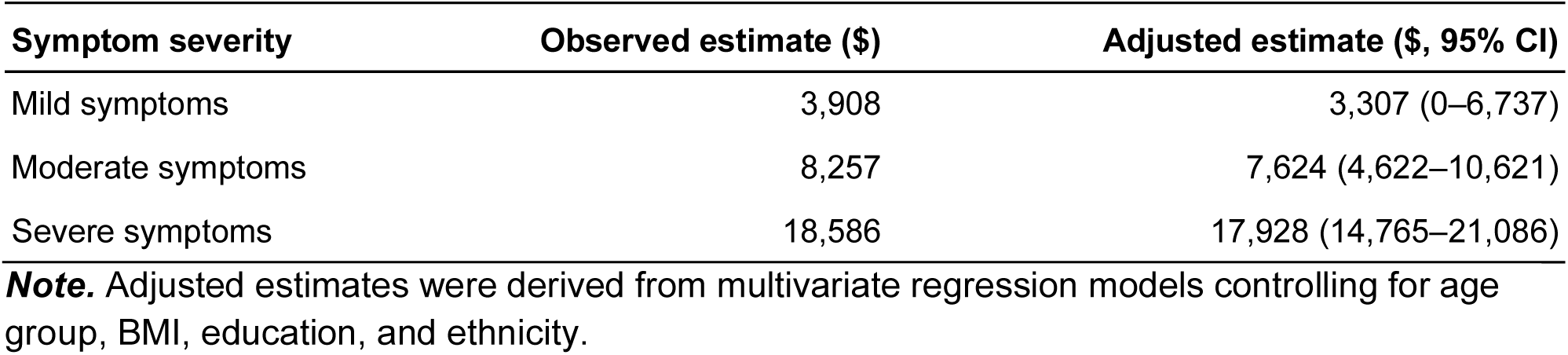
Estimated annual individual productivity loss by symptom severity (observed and adjusted)

**Supplementary Table 6.** Total societal productivity loss of perimenopause.

| Age group | Female population size | Labor force participation rate | Prevalence of perimenopause | Perimenopause-related work impairment* | Median weekly salary (\$USD) | Total weeks per year | Subtotal (\$USD) |
| --- | --- | --- | --- | --- | --- | --- | --- |
| 35 to 39 years | 11,467,859 | 77.40% | 3.11% | 9.77% | \$1,205 | 52 | \$1,687,935,527 |
| 40 to 44 years | 11,115,417 | 77.40% | 24.12% | 9.77% | \$1,205 | 52 | \$12,703,903,140 |
| 45 to 49 years | 10,164,244 | 76.50% | 49.27% | 9.77% | \$1,193 | 52 | \$23,218,458,700 |
| 50 to 54 years | 10,293,667 | 76.50% | 33.48% | 9.77% | \$1,193 | 52 | \$15,981,010,209 |
| 55 to 59 years | 10,307,943 | 60.20% | 8.86% | 9.77% | \$1,097 | 52 | \$3,062,615,468 |
| Total | | | | | | | \$56,653,923,045 |

Note. Perimenopause-related work impairment was estimated based on the observed difference between premenopausal and perimenopausal women

**Supplementary Table 7.**
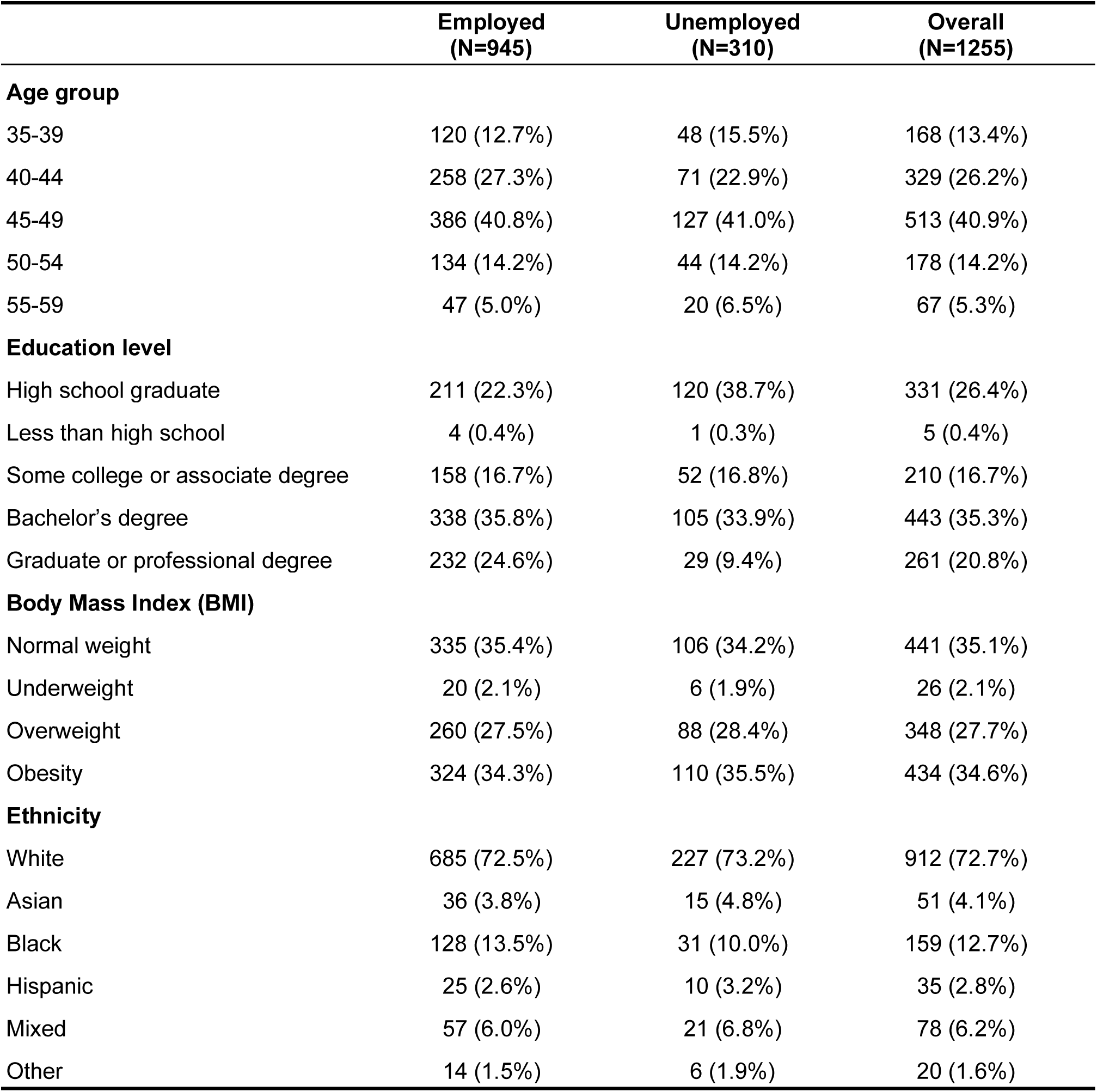
Demographic characteristics of eligible participants by employment status.

|  | **Employed**<br>**(N=945)** | **Unemployed**<br>**(N=310)** | **Overall**<br>**(N=1255)** |
| --- | --- | --- | --- |
| **Age group** |  |  |  |
| 35-39 | 120 (12.7%) | 48 (15.5%) | 168 (13.4%) |
| 40-44 | 258 (27.3%) | 71 (22.9%) | 329 (26.2%) |
| 45-49 | 386 (40.8%) | 127 (41.0%) | 513 (40.9%) |
| 50-54 | 134 (14.2%) | 44 (14.2%) | 178 (14.2%) |
| 55-59 | 47 (5.0%) | 20 (6.5%) | 67 (5.3%) |
| **Education level** |  |  |  |
| High school graduate | 211 (22.3%) | 120 (38.7%) | 331 (26.4%) |
| Less than high school | 4 (0.4%) | 1 (0.3%) | 5 (0.4%) |
| Some college or associate degree | 158 (16.7%) | 52 (16.8%) | 210 (16.7%) |
| Bachelor's degree | 338 (35.8%) | 105 (33.9%) | 443 (35.3%) |
| Graduate or professional degree | 232 (24.6%) | 29 (9.4%) | 261 (20.8%) |
| **Body Mass Index (BMI)** |  |  |  |
| Normal weight | 335 (35.4%) | 106 (34.2%) | 441 (35.1%) |
| Underweight | 20 (2.1%) | 6 (1.9%) | 26 (2.1%) |
| Overweight | 260 (27.5%) | 88 (28.4%) | 348 (27.7%) |
| Obesity | 324 (34.3%) | 110 (35.5%) | 434 (34.6%) |
| **Ethnicity** |  |  |  |
| White | 685 (72.5%) | 227 (73.2%) | 912 (72.7%) |
| Asian | 36 (3.8%) | 15 (4.8%) | 51 (4.1%) |
| Black | 128 (13.5%) | 31 (10.0%) | 159 (12.7%) |
| Hispanic | 25 (2.6%) | 10 (3.2%) | 35 (2.8%) |
| Mixed | 57 (6.0%) | 21 (6.8%) | 78 (6.2%) |
| Other | 14 (1.5%) | 6 (1.9%) | 20 (1.6%) |

### Supplementary Material A. Self-reported reproductive stage question

Below is the self-report question used to classify reproductive stage. Participants were presented with a standardized definition of premenopause, perimenopause, and postmenopause (with visual aid), and asked to select the stage that best described their current experience. This approach facilitated consistent classification based on cycle changes and symptom presentation, particularly among those without a confirmed menopause date.

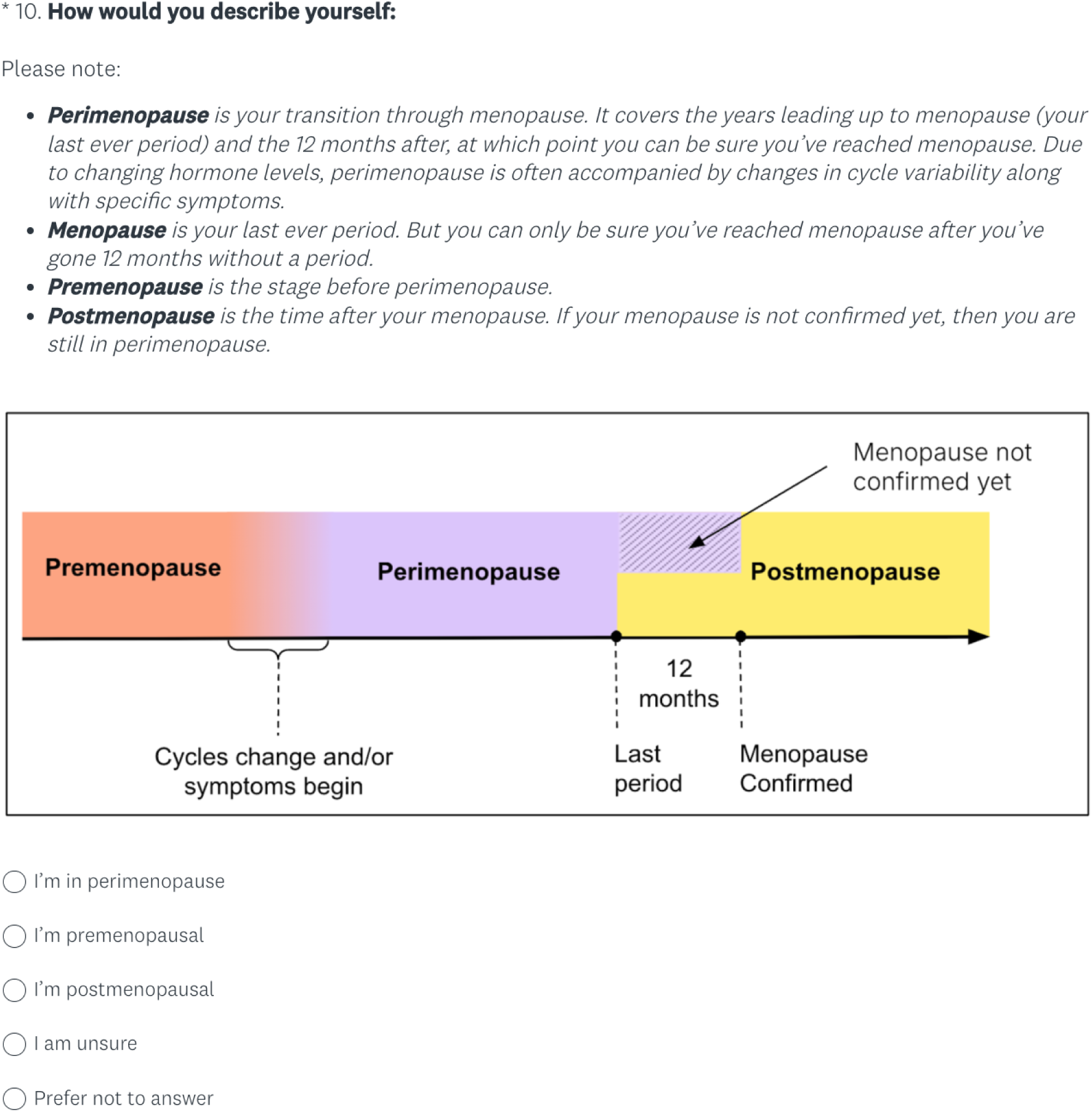

